# Psychometric Properties of the UCSF Fein MAC Educational & Developmental History Questionnaire: A Novel Screening Tool for Capturing Early Life Learning Profiles Across Healthy Aging and Dementia Populations

**DOI:** 10.64898/2025.11.30.25341313

**Authors:** Ezra Mauer, Isabel E. Allen, Rian Bogley, Ryan Newbury, Valentina Diaz, Kaitlin B. Casaletto, Maxime Montembeault, Katherine P. Rankin, Renaud La Joie, Jacob Ziontz, William J. Jagust, Gil D. Rabinovici, Howard J. Rosen, Joel H. Kramer, Bruce L. Miller, Maria Luisa Gorno-Tempini, Zachary A. Miller

## Abstract

**Objective:** Increasing evidence suggests that neurodevelopmental differences substantially alter the expression and course of later-life neurodegenerative diseases. Standard approaches for determining early-life neurodevelopmental differences in aging populations rely largely on chart-based reviews, which poses a methodological challenge due to the varied quality and completeness of medical records. To overcome this limitation, we created the novel Educational & Developmental History (EDevHx) form, a retrospective questionnaire designed to systematically capture early developmental features. Here, we evaluated its psychometric properties among a large sample of cognitively unimpaired, aging adults.

**Methods:** The EDevHx was completed by 677 clinically normal adults aged 46-95 years who underwent standard evaluations to establish their cognitively healthy status.

**Results:** EDevHx items grouped into hypothesized domains (Language, Motor, Visuospatial/Mathematical, Attention, Social) significantly loaded onto their associated domains via confirmatory factor analysis. For each factor, the associated items significantly related to the factor while holding other items constant, indicating a lack of redundancy. Multidimensional scaling analysis showed items were visually grouped within hypothesized domains. Each factor demonstrated acceptable internal consistency. Test-retest reliability ranged from moderate to good, except for the Social factor’s, which was poor. Each factor (as well as two items theorized not to map onto any hypothesized domain) demonstrated convergent/divergent validity with validated questionnaires/neurocognitive tests.

**Conclusion:** The EDevHx tool represents an easily scalable and robust method for capturing early developmental features among aging populations. The present study demonstrates the psychometric adequacy of the EDevHx as a screening instrument, supporting its immediate integration into clinical and research practices alike.

## 1 Introduction

Neurodevelopmental differences, such as non-right-handedness (nRH) and learning disorders (LD), are emerging as critical factors in shaping an individual’s brain health and susceptibility to neurodegenerative diseases (Carr et al., 2024; Miller et al., 2013, 2018, 2025; Rogalski et al., 2008; Siguier et al., 2024). Early observations of elevated nRH in young-onset, language-predominant Alzheimer’s disease led to speculation that neurodevelopmental differences might be associated with later-life degenerative disorders (Geschwind & Galaburda, 1985c; Seltzer & Sherwin, 1983). Increased prevalence of LD was subsequently identified among individuals with language-based neurodegenerative diseases (Rogalski et al., 2008). Further research revealed elevated rates of developmental dyslexia in logopenic variant primary progressive aphasia and increased nRH prevalence in semantic dementia, leading to the hypothesis that early developmental differences might differentially shape the vulnerability of neuroanatomical language networks toward neurodegenerative disease (Miller et al., 2013). Findings of higher rates of mathematical and visuospatial LDs within posterior cortical atrophy further expanded this theorized relationship beyond the language network, producing the hypothesis that domain-specific LDs might be associated with domain-specific neurodegenerative diseases (Miller et al., 2018). More recently, both nRH and LD histories were linked to an earlier onset and impacted the phenotypical targeting of Alzheimer’s disease, supporting the role of neurodevelopmental differences as potential risk factors for dementia (Miller et al., 2025). Lastly, identified genetic correlates of nRH have shown associations with both neurodevelopmental and neurodegenerative disorders (Wiberg et al., 2019). Similarly, the genetics of dyslexia and attention-deficit/hyperactivity disorder have demonstrated links to Alzheimer’s disease (Leffa et al., 2023; Zhu et al., 2024). Beyond neurodegenerative disorders, neurodevelopmental differences also have wide-ranging impacts on quality of life, mental health, occupational success, legal outcomes, as well as life expectancy (Boetsch et al., 1996; Goodman, 2014; Herrera-Araujo et al., 2017; Lollini, 2018; Pullen, 2016; Smith-Spark et al., 2022; Thornton, 2019). Thus, awareness of an individual’s early-life developmental history now takes on much greater significance in aging populations than merely documenting past scholastic-based personal challenges. Without easily accessible information regarding research participants’ neurodevelopment, researchers investigating the associations between developmental differences and adulthood outcomes have had to largely rely on chart review to infer neurodevelopmental history (Siguier et al., 2024). However, the reliability and validity of chart review depend heavily on the quality and features of the medical record (Siems et al., 2020), which presents a significant methodological limitation and impedes progress in this area. This field-wide limitation inspired the creation of a broad-range, questionnaire-based screening tool to systematically capture developmental histories across aging populations.

The present study describes the development and validation of the University of California, San Francisco (UCSF) Fein Memory and Aging Center (MAC) Educational and Developmental History Form (EDevHx), a retrospective questionnaire designed to capture early developmental features. Among a large sample of cognitively unimpaired, aging adults, we examined the factor structure, reliability, and validity of EDevHx responses. Based on lessons learned from this initial pilot study, we also outline plans for the next version of the questionnaire.

## 2 Methods

### 2.1 Scale development

A series of developmental history questions was generated by the study team based on clinical observation and literature review regarding potential neurodevelopmental influences on the presentation and course of neurodegenerative disease. Initial item content was revised by consensus of the study team experts (behavioral neurology and neuropsychology), resulting in a total of 36 questions to be included in the EDevHx. For this study, we focused on the Likert scale questions (items 3-23) that comprised the neurodevelopmental profile. Each question required respondents to rate the degree to which they experienced/displayed a particular difficulty “as a child (prior to age 18)”. For example, the item pertaining to reading asked: “Did you have difficulties reading? (e.g., Were you slower at reading than your peers? Would you need to sound out words to read them? Did school teachers comment on problems with reading or dyslexia?” A 4-point Likert scale was assigned to each item (0 = *never*, 3 = *always*), with higher scores reflecting an endorsement of greater developmental difficulty. Descriptive statistics for EDevHx item responses are presented in **Table 1**. The full EDevHx questionnaire is displayed in the **Supplemental Material**. Though not the focus of the present analysis, the study team also included several dichotomous and nominal items asking about participant hand preference, hand preference among participant family members, history of formal diagnosis of developmental disorders, history of informal suspicion of developmental disorders, and other aspects of past medical and family history.

**Table 1.** Descriptive statistics for EDevHx responses.

| Variable | Minimum | Maximum | Mean | *SD* |
| --- | --- | --- | --- | --- |
| Reading Difficulties | 0.00 | 3.00 | 0.14 | 0.48 |
| Spelling Difficulties | 0.00 | 3.00 | 0.27 | 0.71 |
| Stutter/Stammer | 0.00 | 3.00 | 0.06 | 0.29 |
| Verbal Instructions | 0.00 | 2.00 | 0.13 | 0.38 |
| Writing/Shoelace Difficulties | 0.00 | 3.00 | 0.26 | 0.62 |
| Clumsy | 0.00 | 3.00 | 0.21 | 0.47 |
| Motor Tics | 0.00 | 3.00 | 0.05 | 0.30 |
| Math/Arithmetic Difficulties | 0.00 | 3.00 | 0.40 | 0.72 |
| Geometry Difficulties | 0.00 | 3.00 | 0.42 | 0.74 |
| Left/Right Confusion | 0.00 | 3.00 | 0.21 | 0.58 |
| Navigation Difficulties | 0.00 | 3.00 | 0.28 | 0.62 |
| Difficulties Reading a Map | 0.00 | 3.00 | 0.21 | 0.51 |
| Drawing Difficulties | 0.00 | 3.00 | 0.44 | 0.72 |
| Rhythm Difficulties | 0.00 | 3.00 | 0.28 | 0.59 |
| Frequent Careless Mistakes | 0.00 | 3.00 | 0.38 | 0.56 |
| Bored Easily | 0.00 | 3.00 | 0.55 | 0.67 |
| Attention Difficulties | 0.00 | 3.00 | 0.50 | 0.66 |
| Afraid to Meet Others | 0.00 | 3.00 | 0.50 | 0.68 |
| Prefer Time Alone | 0.00 | 3.00 | 0.82 | 0.70 |
| Afraid to Try New Things | 0.00 | 3.00 | 0.40 | 0.60 |
| Short Temper | 0.00 | 3.00 | 0.27 | 0.49 |

*Note.* EDevHx = Educational and Developmental History Questionnaire, *SD* = standard deviation.

### 2.2 Participants

The EDevHx was piloted on a sample of 677 clinically normal adults (age = 46-95 years, *M _age_* = 69.14, 59.0% female) drawn from two cohorts: the UCSF Fein MAC study participants (*n* = 456) and the University of California, Berkeley Aging Cohort Study (BACS) participants (*n* = 221). Participants from the Fein MAC cohort were classified as clinically normal per consensus multidisciplinary case conference and were deemed functionally intact based on structured interview with a study partner (Clinical Dementia Rating = 0). Participants from the BACS cohort were classified as clinically normal per the following criteria: Mini-Mental State Examination (Folstein et al., 1975) ≥ 25, no serious neurological, psychiatric, or medical illness, and independent community living status. All participants completed the EDevHx via self-report. Detailed demographic characteristics for the full sample (as well as each cohort individually) are presented in **Table 2**.

**Table 2.** Descriptive statistics for demographic characteristics.

|  | Full Sample | Fein MAC<br>(*N* = 456) | BACS<br>(*N* = 221) |
| --- | --- | --- | --- |
| Age, mean (*SD*) | 69.14 (8.33) | 67.29 (8.42) | 72.96 (6.71) |
| Years of Education, mean (*SD*) | 17.35 (2.15) | 17.38 (2.04) | 17.28 (2.37) |
| MMSE, mean (*SD*) | 29.08 (1.15) | 29.15 (1.12) | 28.93 (1.18) |
| **Sex** |  |  |  |
| Female, % | 59.0% | 59.6% | 57.7% |
| Male, % | 41.0% | 40.4% | 42.3% |
| **Handedness** |  |  |  |
| Right-Handed, % | 88.7% | 88.7% | 88.7% |
| Non-Right-Handed, % | 11.3% | 11.3% | 11.3% |
| **Race/Ethnicity** |  |  |  |
| Non-Hispanic White, % | 83.8% | 82.0% | 87.3% |
| Black/African American, % | 2.4% | 1.8% | 3.6% |
| Asian American, % | 7.4% | 9.6% | 2.7% |
| Hispanic/Latino, % | 3.2% | 3.3% | 3.2% |
| Native American, % | 0.6% | 0.7% | 0.5% |
| Unknown/Other, % | 2.7% | 2.6% | 2.7% |

*Note.* MAC = Memory and Aging Center, BACS = Berkeley Aging Cohort Study, *SD* = standard deviation, MMSE = Mini-Mental State Examination.

### 2.3 Additional task-based and questionnaire-based assessment

For testing the construct validity of the EDevHx, participants also completed a series of validated questionnaires and neurocognitive tests. Data from the following questionnaires/tests were used: 1) the Wide Range Achievement Test, Fourth Edition (WRAT4), Word Reading subtest (Wilkinson & Robertson, 2006), 2) the Adult Reading History Questionnaire (ARHQ), a 5-point-Likert-scale (0-4, scale point labels vary by item, lower scores indicate less difficulty) self-report questionnaire assessing past and current reading practices/attitudes (Lefly & Pennington, 2000), 3) Benson Figure copy trial (Possin et al., 2011), a visuospatial construction task, 4) Benton Judgment of Line Orientation (JLO), a visuospatial perception task (Benton et al., 1994), 5) self-reported responses to the Attention subscale of the 4-point-Likert-scale (1 = *rarely/never*, 4 = *almost always/always*) Barratt Impulsiveness Scale (BIS; Patton et al., 1995), 6) the Comprehensive Affect Testing System (CATS) Identity Discrimination subtest, a facial perception task in which participants decide whether neutral faces in a series of face pairings are the same (Froming et al., 2006), and 7) study partner responses on the Anxiety, Angry/Hostility, and Self-Consciousness subscales of the 5-point-Likert-scale (0 = *strongly disagree*, 4 = *strongly agree*) NEO Personality Inventory-3 (NEO-PI-3), which assess proneness to fearfulness/worry, proneness to experience anger, and proneness to shyness/social anxiety respectively (McCrae et al., 2005). Measures were selected to evaluate construct validity by including instruments assessing constructs theoretically aligned with those hypothesized to be reflected in the EDevHx items (to examine convergent validity), as well as instruments assessing conceptually distinct constructs (to examine divergent validity). For the ARHQ, the total number of Likert scale points divided by the total number of points possible (92) was used in analyses. The Likert scale summed scores were used when analyzing data from the remaining questionnaires. For the neurocognitive tests, the total number of correct responses was used in analyses. Due to procedural differences between the Fein MAC and BACS studies, BIS data were only available for the BACS participants while data from the rest of the questionnaires/tests were only available for the Fein MAC participants. To evaluate test-retest reliability, a subset of 109 BACS participants completed the EDevHx on multiple occasions.

### 2.4 Statistical analyses

Following initial grouping of items based on consensus discussion, confirmatory factor analysis (CFA) and a two-dimensional multidimensional scaling analysis were used to examine the dimensionality of the EDevHx. The multidimensional scaling analysis was conducted using standardized item responses so that each item did not skew and bias the similarity structure. To test item redundancy and ensure that the EDevHx was comprised of unique items each providing independent information to the questionnaire, correlation analyses between items was examined and a latent construct analysis was performed. EDevHx subdomain internal consistency was evaluated using the ω coefficient and the Spearman-Brown coefficient. Researchers have recommended use of the ω coefficient over Cronbach’s α because it does not rely on the statistical assumption of tau-equivalence (Jebb et al., 2021). Likewise, the Spearman-Brown coefficient is thought to be most appropriate for two-item scales (Eisinga et al., 2013). For internal consistency, values of .60 or higher were considered acceptable (DeVellis & Thorpe, 2021). Test-retest reliability was evaluated using intraclass correlation and interpreted per the following recommended guidelines: <.50 is poor, .50-.75 is moderate, .75-.90 is good, >.90 is excellent (Koo & Li, 2016). For testing the convergent and divergent validity of the EDevHx with established measures, pairwise correlations were calculated.

### 2.5 Consent statement

The UCSF Committee on Human Research approved all study procedures pertaining to the Fein MAC cohort. The institutional review board at Lawrence Berkeley National Laboratory and the University of California, Berkeley approved all study procedures pertaining to the BACS cohort. All participants provided written informed consent in accordance with the Declaration of Helsinki.

## 3 Results

### 3.1 Dimensionality

To establish EDevHx subscales, the study team first hypothesized the putative cognitive domains onto which each item would map through consensus discussion. The following domains/subscales were generated: Language, Motor, Visuospatial/Mathematical, Attention, and Social. Each item was theorized to map onto one or more of the domains/subscales apart from two items – 1) “Did you find that you were afraid to try new things? (e.g., Did you worry excessively?)” and 2) “Did you have a short temper or throw frequent tantrums? (e.g., Did you get extremely upset if you did not get your way?)” – which were deemed to assess sufficiently independent constructs such that they should not be grouped with any of the domains/subscales. The item pertaining to stuttering/stammering was deemed as plausibly pertaining to either language or motor functioning given its potential conceptualization as a motor speech issue (Namasivayam & van Lieshout, 2011). The item pertaining to left/right confusion was deemed as plausibly pertaining to either motor or visuospatial function because the item’s content does not specify whether the confusion pertains to visuospatial input or motor output. Indeed, the content of the item could plausibly be capturing a Gerstmann-Syndrome-type dysfunction of the angular gyrus (Ardila, 2020), which might suggest the item maps more onto visuospatial functioning. Alternatively, the content of the item could also plausibly be capturing a dyspraxia-type problem, which might suggest the item maps more onto motor functioning. The item pertaining to drawing difficulties was deemed as plausibly pertaining to either motor or visuospatial functioning given the multiple fine motor and visuospatial processing mechanisms that may lead to such difficulties (Senese et al., 2020). Rhythm difficulties was deemed a Motor item based on its localization to the basal ganglia and supplementary motor area (Grahn & Brett, 2007).

Because multiple items were deemed to plausibly map onto more than one domain, an initial CFA was conducted to guide decision-making as to which single domain each item should be grouped with for the sake of parsimoniousness (see **Figure 1**). Analyses were performed using participants with no missing data and also with regression-imputed data for missing values. An analysis of missing data showed the data to be largely missing-at-random. Exceptions were when individual variables were only available in one of the merged datasets. In this first CFA (*χ^2^*(*df* = 139) = 695, *p* < .001, CFI = .813, TLI = .848, and RMSEA = .061) latent Language, Motor, Visuospatial/Mathematical, Attention, and Social factors were indicated by the EDevHx items with 1) the stutter/stammer item loading onto both the Language and Motor factors, 2) the left/right confusion item loading onto both the Motor and Visuospatial/Mathematical factors, and 3) the drawing difficulties item loading onto both the Motor and Visuospatial/Mathematical factors. The stutter/stammer item significantly loaded onto the Motor factor (β = .17, *p* = .025) but not the Language factor (β = .10, *p* = .179), suggesting that the item would be more optimally grouped within the Motor subscale in subsequent analyses. Likewise, the left/right confusion and drawing difficulties items had higher loadings on the Motor factor (βs = .43 and .33, *p*s < .001) compared to the Visuospatial/Mathematical factor (βs = .20 and .30, *p*s < .001), suggesting they should be more optimally grouped within the Motor subscale. All other items significantly loaded onto their hypothesized latent factors.

**Figure 1.**
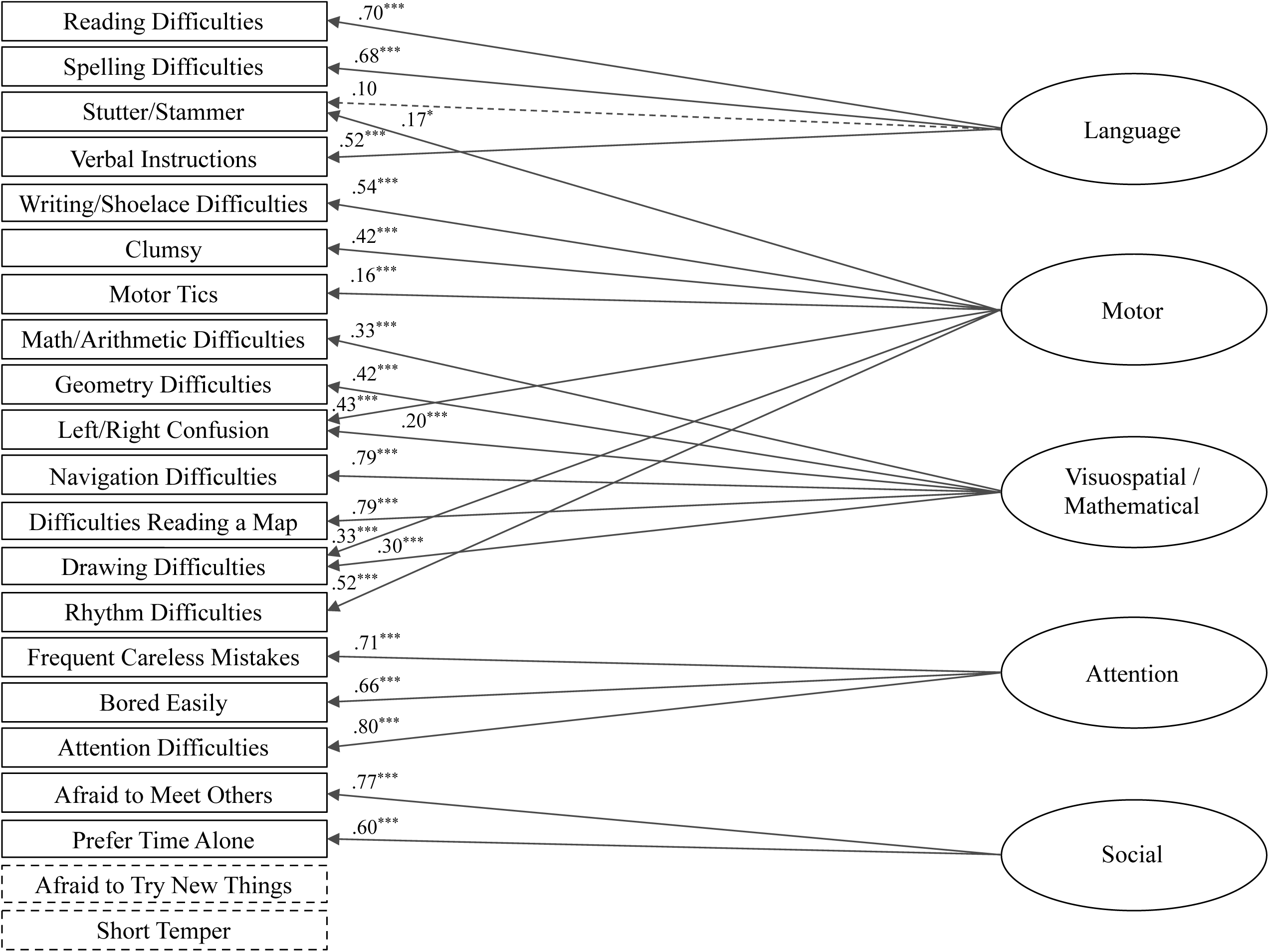
The initial confirmatory factor analysis conducted to evaluate EDevHx factor structure. Items are displayed in the order they appear in the EDevHx. The items “Afraid to Try New Things” and “Short Temper” were not included in the model but are shown for completeness. The numbers in the figures are standardized factor loadings. Nonsignificant paths are displayed with dashed lines. \**p* < .05, \*\**p* < .01, \*\*\**p* < .001.

A second CFA was conducted in which each EDevHx item loaded onto a single putative domain (see **Figure 2**). The second CFA yielded an improved model fit compared to the initial CFA, *χ^2^*(*df* = 142) = 712, *p* < .001, CFI = .881, TLI = .762, and RMSEA = .052. All items significantly loaded onto their hypothesized latent factors. Results of the second CFA suggested it represented an appropriate and parsimonious grouping of EDevHx items. Thus, the second CFA’s item groupings were treated as EDevHx questionnaire subscales in subsequent analyses.

**Figure 2.**
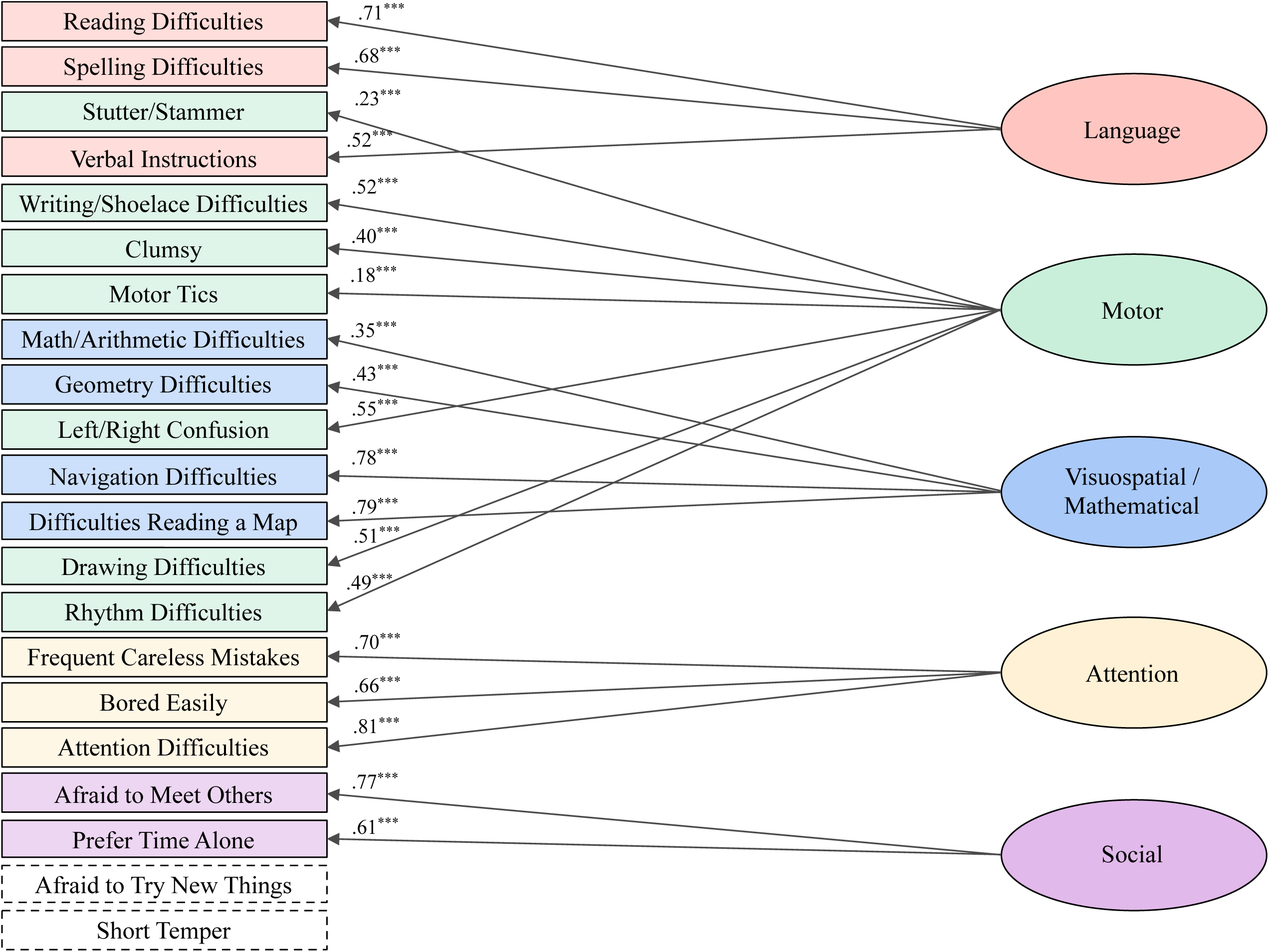
The final confirmatory factor analysis conducted to evaluate EDevHx factor structure. Items are displayed in the order they appear in the EDevHx. The items “Afraid to Try New Things” and “Short Temper” were not included in the model but are shown for completeness. Latent domains are color-coded for clarity. The numbers in the figures are standardized factor loadings. \**p* < .05, \*\**p* < .01, \*\*\**p* < .001.

A redundancy analysis was conducted in which we examined for each factor whether a given factor’s items still significantly related to the factor, holding all other items constant. The results showed that all items significantly related to their respective factors (*p*s < .001), suggesting that no items were redundant.

A two-dimensional multidimensional scaling analysis using standardized item responses was performed to further evaluate the visual similarities and differences of the EDevHx questions (see **Figure 3**). Visual inspection of the results indicated that the items that would be expected to be visually grouped together based upon a priori theories regarding item content and the results of CFA analyses by-and-large were located close to one another within a two-dimensional plane.

**Figure 3.**
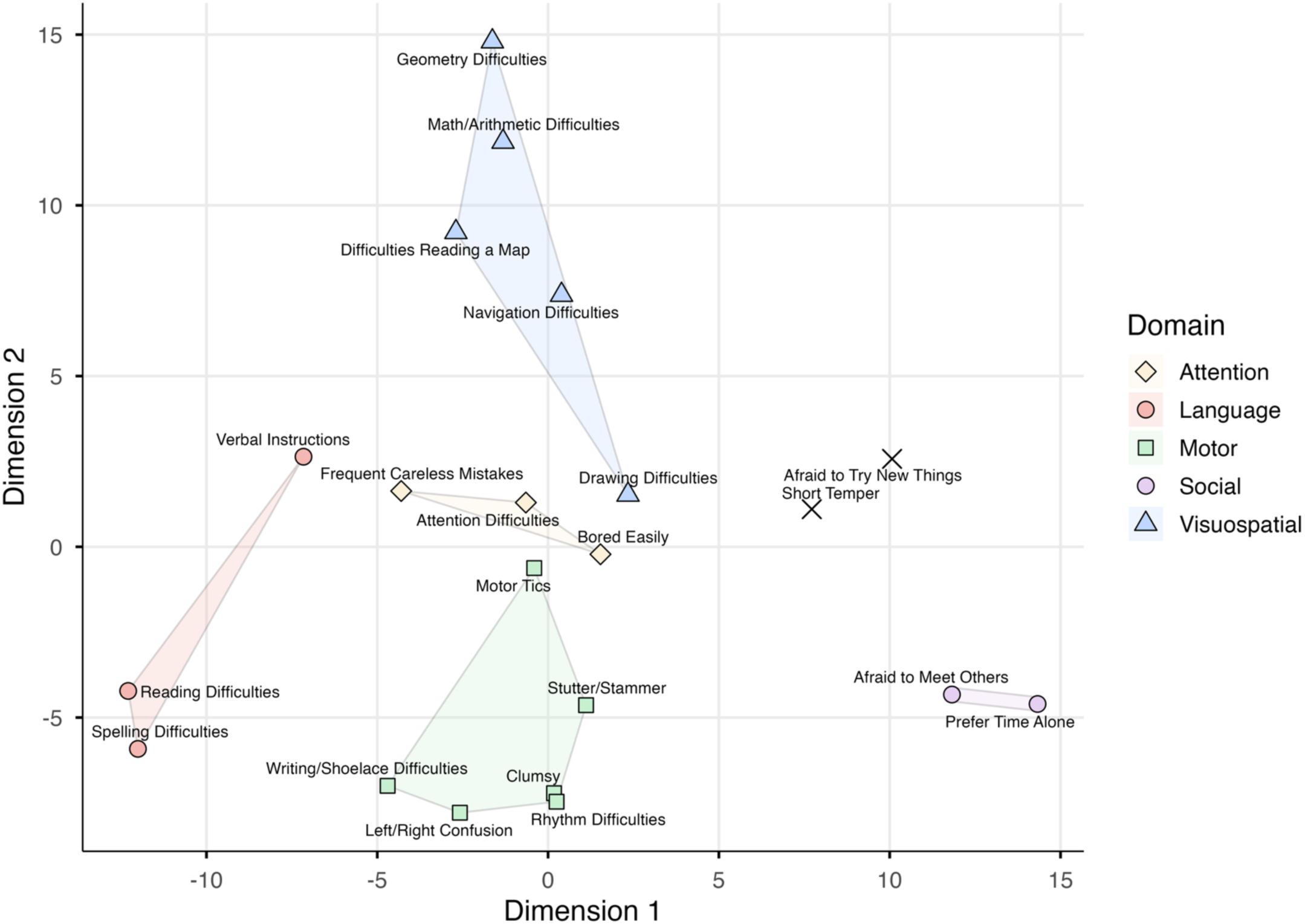
Two-dimensional multidimensional scaling solution illustrating the similarity structure among standardized EDevHx item responses. Items are color-coded according to their hypothesized domains (Language, Motor, Visuospatial/Mathematical, Attention, Social). Spatial proximity reflects similarity in response patterns across participants. Lightly shaded regions indicate convex hulls for domains containing three or more items, and the narrow capsule highlights the spatial relationship between the two items comprising the Social domain.

### 3.2 Reliability

EDevHx subscale internal consistency was tested by calculating the ω coefficient for the Language, Motor, Visuospatial/Mathematical, and Attention subscales and the Spearman-Brown coefficient for the two-item Social subscale. The ω coefficient was .68 for the Language subscale, .61 for the Motor subscale, .81 for the Visuospatial/Mathematical subscale, and .77 for the Attention subscale. Spearman-Brown coefficient was .64 for the Social subscale. Therefore, the internal consistency of all EDevHx subscales was acceptable per DeVellis & Thorpe’s (2021) guidelines.

Test-retest reliability was evaluated by calculating intraclass correlations for each of the EDevHx subscales as well as for the two items that were theorized not to map onto any of the subscales among the subset of 117 BACS participants who completed the questionnaire on multiple occasions. The mean interval between the first and second completion of the EDevHx among this participant subset was 40.09 months (*SD* = 16.82). Intraclass correlation results were as follows: Language subscale = .84, Motor subscale = .89, Visuospatial/Mathematical subscale = .78, Attention subscale = .70, Social subscale = .47, afraid to try new things item = .70, and short temper item = .54. Thus, per interpretive guidelines (Koo & Li, 2016), test-retest reliability ranged from moderate to good, except for that of the Social factor, which was poor.

### 3.3 Validity

Convergent and divergent validity was evaluated by calculating Pearson’s correlations between EDevHx subscale mean scores (and the two non-subscale items’ Likert scale scores) and validated questionnaire/test scores. Descriptive statistics for EDevHx subscale mean scores and the questionnaires/tests used to evaluate construct validity are presented in **Table 3**. Correlations were restricted to theoretically specified convergent and divergent measures for each EDevHx domain rather than examining all possible pairwise associations. Regarding convergent validity, reports of greater language difficulty on the EDevHx was significantly associated with reports of greater reading difficulty on the ARHQ (*r* = .66, *p* < .001) in addition to worse performance on the WRAT4 Word Reading subtest (*r* = -.25, *p* < .001). Greater motor difficulty as reported on the EDevHx was significantly related to worse performance on the Benson Figure copy trial (*r* = -.13, *p* = .008). Reports of greater visuospatial/mathematical difficulty on the EDevHx were significantly associated with worse JLO performance (*r* = -.39, *p* = .001). Attention difficulty reported on the relevant EDevHx subscale and BIS Attention subscale were positively correlated (*r* = .23, *p* = .014). Additionally, the EDevHx Social subscale, afraid to try new things item, and short temper item were positively correlated respectively with the NEO-PI-3 Self-Consciousness (*r* = .33, *p* = .029), Anxiety (*r* = .53, *p* < .001), and Angry/Hostility (*r* = .39, *p* = .008) subscales. Regarding divergent validity, the EDevHx Language and Attention subscales were unrelated to CATS Identity Discrimination performance (*r*s = -.10 and -.12, *p*s = .233 and .156). The EDevHx Motor and Visuospatial/Mathematical subscales were unrelated to WRAT4 Word Reading performance (*r*s = -.06 and -.09, *p*s = .248 and .098). The EDevHx Social subscale, afraid to try new things item, and short temper item were unrelated to Benson Figure copy trial performance (*r*s = .00 to .05, *p*s = .306 to .939). Taken together, results indicated that the EDevHx demonstrated convergent and divergent validity.

**Table 3.** Descriptive statistics for EDevHx subscale mean scores and additional questionnaires/tests used to test construct validity.

| Variable | Minimum | Maximum | Mean | *SD* |
| --- | --- | --- | --- | --- |
| EDevHx Language Subscale | 0.00 | 2.33 | 0.18 | 0.41 |
| EDevHx Motor Subscale | 0.00 | 2.00 | 0.18 | 0.27 |
| EDevHx Visuospatial/Mathematical Subscale | 0.00 | 2.80 | 0.36 | 0.45 |
| EDevHx Attention Subscale | 0.00 | 2.67 | 0.48 | 0.52 |
| EDevHx Social Subscale | 0.00 | 3.00 | 0.63 | 0.61 |
| WRAT4 Word Reading | 53.00 | 70.00 | 64.69 | 3.03 |
| ARHQ | 0.00 | 0.76 | 0.16 | 0.11 |
| Benson Figure Copy | 10.00 | 17.00 | 15.36 | 0.98 |
| JLO | 8.00 | 15.00 | 13.07 | 1.74 |
| BIS Attention Subscale | 4.00 | 14.00 | 8.79 | 2.21 |
| CATS Identity Discrimination | 9.00 | 12.00 | 11.78 | 0.55 |
| NEO-PI-3 Anxiety Subscale | 8.00 | 32.00 | 17.96 | 5.09 |
| NEO-PI-3 Angry/Hostility Subscale | 8.00 | 34.00 | 16.87 | 5.45 |
| NEO-PI-3 Self-Consciousness Subscale | 9.00 | 28.00 | 16.11 | 4.57 |

*Note.* EDevHx = Educational and Developmental History Questionnaire, *SD* = standard deviation, WRAT4 = Wide Range Achievement Test, Fourth Edition, ARHQ = Adult Reading History Questionnaire, JLO = Benton Judgment of Line Orientation, BIS = Barratt Impulsiveness Scale, CATS = Comprehensive Affect Testing System, NEO-PI-3 = NEO Personality Inventory-3.

## 4 Discussion

Neurodevelopmental differences, historically unacknowledged in later life health and function, have been increasingly recognized for their influence on quality of life, mental health, occupational success, legal outcomes, and life expectancy (Boetsch et al., 1996; Goodman, 2014; Herrera-Araujo et al., 2017; Lollini, 2018; Pullen, 2016; Smith-Spark et al., 2022; Thornton, 2019). More recently, neurodevelopmental differences have been recognized for their role in shaping the expression/phenotypical targeting and timing of neurodegenerative disease onset (Carr et al., 2024; Miller et al., 2013, 2018, 2025; Siguier et al., 2024). With this growing body of evidence, there is urgent need to scale up and standardize the collection of neurodevelopmental data across healthy aging and dementia populations. However, obtaining accurate developmental histories within the aging population presents unique challenges. These challenges have been primarily structural – due to cultural practices such as forced right-handedness (McManus et al., 2010) and a historical lack of awareness of developmental differences (Agrawal et al., 2019; Pullen, 2016) – and persist across many regions of the globe (Mather et al., 2020). As a result, many older adults grew up before neurodevelopmental diagnostic assessments became widely available. Even as assessments for neurodevelopmental differences have become more accessible, they have largely been limited to school-aged populations (Agrawal et al., 2019; Pullen, 2016). Consequently, detailed neurodevelopmental information about aging research participants has remained largely inaccessible.

To address this gap, researchers – including ourselves – have often relied on retrospective chart-based studies to infer developmental histories (Siguier et al., 2024). While chart-based reviews can provide valuable insights, they are logistically challenging and prone to methodological limitations due to the variability in the quality and completeness of medical records (Siems et al., 2020). By contrast, questionnaires offer significant advantages, including standardization, scalability, and ease of deployment (Ranganathan & Caduff, 2023). Drawing on our clinical observations and literature documenting potential neurodevelopmental influences on the presentation and progression of neurodegenerative diseases, incorporating lessons learned from the field of cognitive behavioral neurology and neuropsychology, we developed the UCSF Fein MAC EDevHx screening tool, a retrospective questionnaire designed to capture early educational and developmental history across a wide range of cognitive domains and behaviors.

The goal of the EDevHx form was to create a brief screening tool with equal clinical and research application that demonstrates cognitive, behavioral, and neuroanatomical relevance. Unlike current tools created to capture history of early life learning differences in adults, such as the ARHQ and the 10-item short form of the Autism Spectrum Quotient questionnaire (Allison et al., 2012; Lefly & Pennington, 2000), the EDevHx form was designed specifically to be transdiagnostic – with the goal of capturing shared mechanisms across various neurodevelopmental diagnostic labels. To that end, the Likert scale items evaluated in the present study were designed as open-ended inquiries into learning history and performance across cognitive domains. Thus, these items, provide opportunities to investigate the proportion of individuals, for instance, who endorse reading and mathematical difficulties. Among these Likert scale items, we purposefully limited the use of diagnostic labels or medical jargon so as to reflect an agnostic inquiry into an individual’s academic performance up through high school. With this, an individual would be able to rate their perceived ability in a particular area of inquiry, like reading, without needing to rely on diagnoses that may have been inaccessible. Additionally, current LD diagnostic labels are largely based on cutoffs that establish healthy-versus-impaired performances, despite the reality that scholastic abilities fall along a broad spectrum from impaired to superior. Given this, we designed the EDevHx to capture behaviors ranging from normal to impaired. Together, this approach allowed us to generate an individualized neurodevelopmental profile; in the present study, we focused our analyses on the portion of the EDevHx used to generate these individualized neurodevelopmental cognitive profiles.

We piloted the EDevHx among a large group of 677 cognitively unimpaired, aging adults participating in studies at the UCSF Fein MAC and the University of California, BACS. We examined the factor structure, reliability, and validity of participant responses within the EDevHx’s neurodevelopmental cognitive profile section (items 3-23). CFA and multidimensional scaling analysis supported the hypothesized grouping of items into five subscales: Language, Motor, Visuospatial/Mathematical, Attention, and Social. Two items (“Did you find that you were afraid to try new things?” and “Did you have a short temper or throw frequent tantrums?”) were deemed to assess sufficiently independent constructs such that they were not grouped with any of the subscales. Although CFI/TLI values were modest in the final CFA, RMSEA indicated good absolute fit. Importantly, several EDevHx items exhibited highly skewed response distributions with strong floor effects in this cognitively healthy sample, a pattern that can attenuate incremental fit indices such as CFI and TLI in CFA models. Given the screening purpose of the instrument and the theoretical coherence of factor loadings, we interpreted the final CFA as providing acceptable structural support. Redundancy analysis found no evidence of item redundancy. Regarding reliability, all subscales demonstrated acceptable internal consistency. Because the EDevHx was intentionally designed as a brief screening instrument rather than a comprehensive diagnostic measure, moderate internal consistency values were anticipated and considered acceptable for initial validation. Test-retest reliability of the subscales and the two independent items ranged from moderate to good, apart from that of the Social subscale, which was poor. Evaluation of construct validity using a series of previously validated questionnaires and neurocognitive tests showed that the subscales and two independent items each demonstrated convergent and divergent validity. Regarding convergent validity, the Language subscale was correlated with worse word reading performance (WRAT4 Word Reading) in addition to self-reported difficulty on a reading practices/attitudes questionnaire (ARHQ). The Motor subscale was associated with worse performance on a visuospatial construction task (Benson Figure copy trial). Participant responses on the Visuospatial/Mathematical subscale were associated with lower visuospatial perception task performance (JLO). The Attention subscale was positively correlated with self-reported questionnaire ratings of attentional difficulty (BIS Attention subscale). The Social subscale, afraid to try new things item, and short temper item were positively correlated with study-partner questionnaire ratings of shyness/social anxiety (NEO-PI-3 Self-Consciousness subscale), fearfulness/worry (NEO-PI-3 Anxiety subscale), and anger (NEO-PI-3 Angry/Hostility subscale) respectively. Regarding divergent validity, the Language and Attention subscales were unrelated to performance on a facial perception task (CATS Identity Discrimination). The Motor and Visuospatial/Mathematical subscales were unrelated to word reading performance (WRAT4 Word Reading). Likewise, the Social subscale, afraid to try new things item, and short temper item were unrelated to performance on a visuospatial construction task (Benson Figure copy trial).

The EDevHx and its validation study have several strengths. First, the large sample size enhances confidence in the findings regarding the questionnaire’s psychometric properties. Second, the EDevHx is easily deployable and scalable in both research and clinical settings, representing a significant improvement over chart review approaches. Third, the flexibility of the EDevHx allows researchers to tailor item groupings to specific research questions. For instance, investigations focused on written language difficulties could exclusively examine items related to reading and spelling with the Language subscale. The piloting and validation of the EDevHx have provided valuable insights into both the utility of the instrument and areas for improvement. While the psychometric properties of the EDevHx were generally acceptable for a screening instrument, the Social subscale demonstrated poor test-retest reliability. Future versions of the questionnaire will aim to improve this subscale’s reliability. Expanding the questionnaire to better capture impulsivity and hyperactivity, key symptoms of attention-deficit/hyperactivity disorder, should be considered in future versions. Additionally, as our research group identified links between earlier life systematizing behaviors and frontotemporal dementia (Miller et al., 2020), we realized the need to more specifically assess social communication differences and restricted/repetitive behaviors characteristic of autism spectrum disorder. Furthermore, the questionnaire’s current focus on normal-to-impaired performances (a choice we made in the piloting phase in order to have this tool parallel related questionnaires, which themselves were modeled on the focus within medical research on disability) limits our ability to capture an individual’s strengths and “special talents.” Adjusting the questionnaire to better capture strengths would enrich our understanding of developmental differences considering the literature regarding “hidden strengths” within LD populations (Geschwind & Galaburda, 1985a, 1985b). As we continue to expand the scope and scale of the EDevHx, it remains critical to strike a balance between comprehensiveness and brevity while maintaining or improving psychometric properties, as an essential aspect of the EDevHx is that it remains a screening tool – with the ability to indicate the utility of incorporating more in-depth validated questionnaires focused on specific neurodevelopmental disorders in the context of research/clinical investigations. Finally, in this study, our focus was on psychometric properties, which is why our analysis of neurocognitive associations was limited. With larger data collections and broader access to cognitive tests, we expect to more precisely investigate how our screening tool’s items relate to individual cognitive processes underlying the relevant domains of functioning (e.g., investigating the Language subscale’s relations with phonological awareness rather than just word reading broadly). Likewise, many of the present study’s participants have undergone structural and functional neuroimaging, which allows for future studies exploring the anatomical associations of our EDevHx-based developmental profiles. Thus, it is our expectation that lessons learned from the EDevHx will rapidly advance the understanding of childhood neurodevelopment while providing the first reliable means of studying the natural history of aging across populations with varied developmental differences.

The EDevHx represents an easily scalable method for capturing early developmental features, offering a significant methodological improvement over traditional chart review approaches. While the initial motivation for designing the EDevHx was to scale up collections in elderly populations as a means of more clearly establishing the putative domain-specific associations between developmental differences and select neurodegenerative disorders (Carr et al., 2024; Miller et al., 2013, 2018, 2025; Rogalski et al., 2008; Siguier et al., 2024), its ability to identify developmental differences across large adult populations also provides an opportunity to investigate the relationship between development and a myriad of health and social conditions. At its core, the EDevHx stands to deepen our understanding of the neurobiological basis of neurodevelopmental differences themselves.

## Supporting information

Supplemental Material: EDevHx Questionnaire

## Data Availability

All data produced in the present study are available upon reasonable request to the authors.

## Acknowledgements

We wish to express our sincere gratitude to the participants in our research program who made this work possible.

## References

Agrawal, J., Barrio, B. L., Kressler, B., Hsiao, Y.-J., & Shankland, R. K. (2019). International Policies, Identification, and Services for Students with Learning Disabilities: An Exploration across 10 Countries. Learning Disabilities: A Contemporary Journal, 17(1), 95–113.

Allison, C., Auyeung, B., & Baron-Cohen, S. (2012). Toward Brief “Red Flags” for Autism Screening: The Short Autism Spectrum Quotient and the Short Quantitative Checklist in 1,000 Cases and 3,000 Controls. Journal of the American Academy of Child & Adolescent Psychiatry, 51(2), 202–212.e7. 10.1016/j.jaac.2011.11.003

Ardila, A. (2020). Gerstmann Syndrome. Current Neurology and Neuroscience Reports, 20(11), 48. 10.1007/s11910-020-01069-9

Benton, A. L., Sivan, A. B., Hamsher, K. de S., Varney, N. R., & Spreen, O. (1994). Contributions to Neuropsychological Assessment: A Clinical Manual. Oxford University Press.

Boetsch, E. A., Green, P. A., & Pennington, B. F. (1996). Psychosocial correlates of dyslexia across the life span. Development and Psychopathology, 8(3), 539–562. 10.1017/S0954579400007264

Carr, R. H., Eom, G. D., & Brown, E. E. (2024). Attention-Deficit/Hyperactivity Disorder as a Potential Risk Factor for Dementia and Other Neurocognitive Disorders: A Systematic Review. Journal of Alzheimer’s Disease, 98(3), 773–792. 10.3233/JAD-230904

DeVellis, R. F., & Thorpe, C. T. (2021). Scale Development: Theory and Applications. SAGE Publications.

Eisinga, R., Grotenhuis, M. te, & Pelzer, B. (2013). The reliability of a two-item scale: Pearson, Cronbach, or Spearman-Brown? International Journal of Public Health, 58(4), 637–642. 10.1007/s00038-012-0416-3

Folstein, M. F., Folstein, S. E., & McHugh, P. R. (1975). “Mini-mental state”: A practical method for grading the cognitive state of patients for the clinician. Journal of Psychiatric Research, 12(3), 189–198. 10.1016/0022-3956(75)90026-6

Froming, K. B., Ekman, P., & Levy, M. (2006). Comprehensive Affect Testing System. Psychology Software, Inc. https://doi.apa.org/doi/10.1037/t06823-000

Geschwind, N., & Galaburda, A. M. (1985a). Cerebral Lateralization: Biological Mechanisms, Associations, and Pathology: I. A Hypothesis and a Program for Research. Archives of Neurology, 42(5), 428–459. 10.1001/archneur.1985.04060050026008

Geschwind, N., & Galaburda, A. M. (1985b). Cerebral Lateralization: Biological Mechanisms, Associations, and Pathology: II. A Hypothesis and a Program for Research. Archives of Neurology, 42(6), 521–552. 10.1001/archneur.1985.04060060019009

Geschwind, N., & Galaburda, A. M. (1985c). Cerebral Lateralization: Biological Mechanisms, Associations, and Pathology: III. A Hypothesis and a Program for Research. Archives of Neurology, 42(7), 634–654. 10.1001/archneur.1985.04060070024012

Goodman, J. (2014). The Wages of Sinistrality: Handedness, Brain Structure, and Human Capital Accumulation. Journal of Economic Perspectives, 28(4), 193–212. 10.1257/jep.28.4.193

Grahn, J. A., & Brett, M. (2007). Rhythm and beat perception in motor areas of the brain. Journal of Cognitive Neuroscience, 19(5), 893–906. 10.1162/jocn.2007.19.5.893

Herrera-Araujo, D., Shaywitz, B. A., Holahan, J. M., Marchione, K. E., Michaels, R., Shaywitz, S. E., & Hammitt, J. K. (2017). Evaluating Willingness to Pay as a Measure of the Impact of Dyslexia in Adults. Journal of Benefit-Cost Analysis, 8(1), 24–48. 10.1017/bca.2017.3

Jebb, A. T., Ng, V., & Tay, L. (2021). A Review of Key Likert Scale Development Advances: 1995–2019. Frontiers in Psychology, 12. 10.3389/fpsyg.2021.637547

Koo, T. K., & Li, M. Y. (2016). A Guideline of Selecting and Reporting Intraclass Correlation Coefficients for Reliability Research. Journal of Chiropractic Medicine, 15(2), 155–163. 10.1016/j.jcm.2016.02.012

Leffa, D. T., Ferrari-Souza, J. P., Bellaver, B., Tissot, C., Ferreira, P. C. L., Brum, W. S., Caye, A., Lord, J., Proitsi, P., Martins-Silva, T., Tovo-Rodrigues, L., Tudorascu, D. L., Villemagne, V. L., Cohen, A. D., Lopez, O. L., Klunk, W. E., Karikari, T. K., Rosa-Neto, P., Zimmer, E. R., … Pascoal, T. A. (2023). Genetic risk for attention-deficit/hyperactivity disorder predicts cognitive decline and development of Alzheimer’s disease pathophysiology in cognitively unimpaired older adults. Molecular Psychiatry, 28(3), 1248–1255. 10.1038/s41380-022-01867-2

Lefly, D. L., & Pennington, B. F. (2000). Reliability and validity of the Adult Reading History Questionnaire. Journal of Learning Disabilities, 33(3), 286–296. 10.1177/002221940003300306

Lollini, A. (2018). Brain Equality: Legal Implications of Neurodiversity in a Comparative Perspective. New York University Journal of International Law and Politics, 51, 69.

Mather, N., White, J., & Youman, M. (2020). Dyslexia Around the World: A Snapshot. Learning Disabilities: A Multidisciplinary Journal, 25(1). 10.18666/LDMJ-2020-V25-I1-9552

McCrae, R. R., Costa, Jr., Paul T., & Martin, T. A. (2005). The NEO–PI–3: A More Readable Revised NEO Personality Inventory. Journal of Personality Assessment, 84(3), 261–270. 10.1207/s15327752jpa8403_05

McManus, I. C., Moore, J., Freegard, M., & Rawles, R. (2010). Science in the Making: Right Hand, Left Hand. III: Estimating historical rates of left-handedness. Laterality, 15(1–2), 186–208. 10.1080/13576500802565313

Miller, Z. A., Allen, I. E., Neylan, K. D., Diggs, R. T., Bogely, R. L., Palser, E., Perry, D. C., Brown, J., Sturm, V., Rabinovici, G. D., Rosen, H. J., Kramer, J. H., Rankin, K., Grinberg, L. T., Seeley, W. W., Gorno-Tempini, M., & Miller, B. L. (2020). Divergent effects of education and occupation history on age at onset within Alzheimer’s and frontotemporal dementia. Alzheimer’s & Dementia, 16(S6), e046656. 10.1002/alz.046656

Miller, Z. A., Mandelli, M. L., Rankin, K. P., Henry, M. L., Babiak, M. C., Frazier, D. T., Lobach, I. V., Bettcher, B. M., Wu, T. Q., Rabinovici, G. D., Graff-Radford, N. R., Miller, B. L., & Gorno-Tempini, M. L. (2013). Handedness and language learning disability differentially distribute in progressive aphasia variants. Brain, 136(11), 3461–3473. 10.1093/brain/awt242

Miller, Z. A., Ossenkoppele, R., Graff-Radford, N. R., Allen, I. E., Shwe, W., Rosenberg, L., Olguin, D. J., Erkkinen, M. G., Butler, P. M., Spina, S., Yokoyama, J. S., Desikan, R. S., Scheltens, P., van der Flier, W., Pijnenburg, Y., Wolters, E., Rademakers, R., Geschwind, D. H., Kramer, J. H., … Gorno-Tempini, M. L. (2025). Neurodevelopment and neural environment inform Alzheimer’s disease age at onset and phenotype. Alzheimer’s & Dementia, 21(9), e70668. 10.1002/alz.70668

Miller, Z. A., Rosenberg, L., Santos-Santos, M. A., Stephens, M., Allen, I. E., Hubbard, H. I., Cantwell, A., Mandelli, M. L., Grinberg, L. T., Seeley, W. W., Miller, B. L., Rabinovici, G. D., & Gorno-Tempini, M. L. (2018). Prevalence of mathematical and visuospatial learning disabilities in patients with posterior cortical atrophy. JAMA Neurology, 75(6), 728–737. 10.1001/jamaneurol.2018.0395

Namasivayam, A. K., & van Lieshout, P. (2011). Speech motor skill and stuttering. Journal of Motor Behavior, 43(6), 477–489. 10.1080/00222895.2011.628347

Patton, J. H., Stanford, M. S., & Barratt, E. S. (1995). Factor structure of the Barratt impulsiveness scale. Journal of Clinical Psychology, 51(6), 768–774. 10.1002/1097-4679(199511)51:6%253C768::aid-jclp2270510607%253E3.0.co;2-1

Possin, K. L., Laluz, V. R., Alcantar, O. Z., Miller, B. L., & Kramer, J. H. (2011). Distinct neuroanatomical substrates and cognitive mechanisms of figure copy performance in Alzheimer’s disease and behavioral variant frontotemporal dementia. Neuropsychologia, 49(1), 43–48. 10.1016/j.neuropsychologia.2010.10.026

Pullen, P. C. (2016). Historical and Current Perspectives on Learning Disabilities in the United States. Learning Disabilities - A Contemporary Journal, 14(1), 25–37.

Ranganathan, P., & Caduff, C. (2023). Designing and validating a research questionnaire—Part 1. Perspectives in Clinical Research, 14(3), 152. 10.4103/picr.picr_140_23

Rogalski, E., Johnson, N., Weintraub, S., & Mesulam, M. (2008). Increased frequency of learning disability in patients with primary progressive aphasia and their first-degree relatives. Archives of Neurology, 65(2), 244–248. 10.1001/archneurol.2007.34

Seltzer, B., & Sherwin, I. (1983). A Comparison of Clinical Features in Early- and Late-Onset Primary Degenerative Dementia: One Entity or Two? Archives of Neurology, 40(3), 143–146. 10.1001/archneur.1983.04050030037006

Senese, V. P., Zappullo, I., Baiano, C., Zoccolotti, P., Monaco, M., & Conson, M. (2020). Identifying neuropsychological predictors of drawing skills in elementary school children. Child Neuropsychology, 26(3), 345–361. 10.1080/09297049.2019.1651834

Siems, A., Banks, R., Holubkov, R., Meert, K. L., Bauerfeld, C., Beyda, D., Berg, R. A., Bulut, Y., Burd, R. S., Carcillo, J., Dean, J. M., Gradidge, E., Hall, M. W., McQuillen, P. S., Mourani, P. M., Newth, C. J. L., Notterman, D. A., Priestley, M. A., Sapru, A., … Pollack, M. M. (2020). Structured Chart Review: Assessment of a Structured Chart Review Methodology. Hospital Pediatrics, 10(1), 61–69. 10.1542/hpeds.2019-0225

Siguier, P. L. M., Planton, M., Baudou, E., Chaix, Y., Delage, A., Rafiq, M., Wolfrum, M., Gérard, F., Jucla, M., & Pariente, J. (2024). Can neurodevelopmental disorders influence the course of neurodegenerative diseases? A scoping review. Ageing Research Reviews, 99, 102354. 10.1016/j.arr.2024.102354

Smith-Spark, J. H., Gordon, R., & Jansari, A. S. (2022). The impact of developmental dyslexia on workplace cognition: Evidence from a virtual reality environment. Proceedings of the 33rd European Conference on Cognitive Ergonomics, 1–4. 10.1145/3552327.3552340

Thornton, J. (2019). People with learning disabilities have lower life expectancy and cancer screening rates. BMJ, 364, l404. 10.1136/bmj.l404

Wiberg, A., Ng, M., Al Omran, Y., Alfaro-Almagro, F., McCarthy, P., Marchini, J., Bennett, D. L., Smith, S., Douaud, G., & Furniss, D. (2019). Handedness, language areas and neuropsychiatric diseases: Insights from brain imaging and genetics. Brain, 142(10), 2938–2947. 10.1093/brain/awz257

Wilkinson, G. S., & Robertson, G. J. (2006). Wide range achievement test (WRAT4). Psychological Assessment Resources.

Zhu, P., Gao, S., Wu, S., Li, X., Huang, C., Chen, Y., & Liu, G. (2024). Causal relationships between dyslexia and the risk of eight dementias. Translational Psychiatry, 14(1), 1–10. 10.1038/s41398-024-03082-9

