## Supplemental Material: EDevHx Questionnaire for "Psychometric Properties of the UCSF Fein MAC Educational & Developmental History Questionnaire: A Novel Screening Tool for Capturing Early Life Learning Profiles Across Healthy Aging and Dementia Populations"

### UCSF FEIN MEMORY AND AGING CENTER
### EDUCATIONAL AND DEVELOPMENTAL HISTORY FORM

**Demographics**

| | | Left | Ambidextrous | Forced-Right | Right | N/A |
|---|---|---|---|---|---|---|
| 1. | **What is your hand preference?** e.g. Do you consider yourself to be left-handed, ambidextrous, forced-right-handed (were originally left-handed or ambidextrous and forced to become right), or right-handed? | ☐ | ☐ | ☐ | ☐ | ☐ |

| | | Yes | No | I don't know |
|---|---|---|---|---|
| 2. | **Are any family members NOT right handed?** e.g. Were/are any of your parents, siblings, or children left-handed, ambidextrous, and/or forced right-handed? | ☐ | ☐ | ☐ |

☐Mother, ☐Father, ☐Sibling, ☐Child, ☐Other family member:

**Details:**

#### AS A CHILD (PRIOR TO AGE 18):

| | | 0 Never | 1 Sometimes | 2 Often | 3 Always | 4 N/A |
|---|---|---|---|---|---|---|
| 3. | **Did you have difficulties reading?** e.g. Were you slower at reading than your peers? Would you need to sound out words to read them? Did school teachers comment on problems with reading or dyslexia? | ☐ | ☐ | ☐ | ☐ | ☐ |
| 4. | **Did you have difficulties with spelling?** e.g. Did school teachers comment on problems with spelling? | ☐ | ☐ | ☐ | ☐ | ☐ |
| 5. | **Did you stutter or stammer?** | ☐ | ☐ | ☐ | ☐ | ☐ |
| 6. | **Did you have problems comprehending verbal instruction in school?** e.g. Did you have difficulty related specifically to verbal instructions as opposed to written? | ☐ | ☐ | ☐ | ☐ | ☐ |
| 7. | **Did you have difficulty with your handwriting and/or difficulties with tying shoelaces?** e.g. Did parents or school teachers comment on these problems? | ☐ | ☐ | ☐ | ☐ | ☐ |
| 8. | **Did you think you were clumsy?** e.g. Did you break items frequently? Bump into things or other people? Would people comment on you being clumsy? | ☐ | ☐ | ☐ | ☐ | ☐ |
| 9. | **Did you display motor tics?** e.g. Would you frequently clear your throat, or exhibit excessive blinking, and/or head jerking? | ☐ | ☐ | ☐ | ☐ | ☐ |
| 10. | **Did you have difficulties with math/arithmetic?** e.g. Did you require greater assistance mastering mathematical concepts? Did teachers comment on problems with math? | ☐ | ☐ | ☐ | ☐ | ☐ |
| 11. | **Did you have difficulties with geometry?** e.g. Did you have difficulties with relating numerical values to visual concepts? | ☐ | ☐ | ☐ | ☐ | ☐ |
| 12. | **Did you have Left/Right confusion?** e.g. Did you have difficulties telling your left from right? | ☐ | ☐ | ☐ | ☐ | ☐ |
| 13. | **Did you have difficulties with navigation/directions?** e.g. Would you get lost without someone guiding you? | ☐ | ☐ | ☐ | ☐ | ☐ |
| 14. | **Did you have difficulties reading a map?** | ☐ | ☐ | ☐ | ☐ | ☐ |
| 15. | **Did you have difficulties drawing?** e.g. Was it challenging for you to copy images or proportions? | ☐ | ☐ | ☐ | ☐ | ☐ |
| 16. | **Did you have difficulties following a rhythm?** e.g. Did you have difficulties clapping along and keeping in time with the music? | ☐ | ☐ | ☐ | ☐ | ☐ |

| | | | Never | Sometimes | Often | Always | N/A |
|---|---|---|---|---|---|---|---|
| | 17. | **Did you make frequent careless mistakes?** e.g. Did you have problems paying attention to details or instructions? | ☐ | ☐ | ☐ | ☐ | ☐ |
| | 18. | **Did you find that you were bored easily?** e.g. Did you move from one activity to the next? | ☐ | ☐ | ☐ | ☐ | ☐ |
| | 19. | **Did you have difficulty paying attention in school?** e.g. Did you daydream often? Were you constantly lost in your own thoughts? | ☐ | ☐ | ☐ | ☐ | ☐ |
| | 20. | **Did you find you were afraid to meet other people?** e.g. Did you avoid social events on purpose? | ☐ | ☐ | ☐ | ☐ | ☐ |
| | 21. | **Did you prefer spending time alone?** | ☐ | ☐ | ☐ | ☐ | ☐ |
| | 22. | **Did you find that you were afraid to try new things?** e.g. Did you worry excessively? | ☐ | ☐ | ☐ | ☐ | ☐ |
| | 23. | **Did you have a short temper or throw frequent tantrums?** e.g. Did you get extremely upset if you did not get your way? | ☐ | ☐ | ☐ | ☐ | ☐ |

#### AS A CHILD (PRIOR TO AGE 18):

| Past Medical and Family History | | | Yes | No | I don't know |
|---|---|---|---|---|---|
| | 24. | **Was your speech delayed?** e.g. Did your family comment on your late talking compared to other children? | ☐ | ☐ | ☐ |
| | 25. | **Were you ever diagnosed as color blind?** e.g. Did you have difficulties distinguishing similar shades of color? | ☐ | ☐ | ☐ |
| | 26. | **Did you have visual difficulties beyond needing regular glasses or contacts?** e.g. Did you ever have strabismus (cross-eyes) or need eye surgery or patching? | ☐ | ☐ | ☐ |
| | 27. | **Did you have difficulties with depth perception?** e.g. Did you have trouble telling how close or far objects were relative to you? | ☐ | ☐ | ☐ |
| | 28. | **Did you have trouble recognizing faces?** e.g. Did faces all look the same? | ☐ | ☐ | ☐ |
| | 29. | **Did you ever suffer from a seizure, febrile seizure, or epileptic fit?** e.g. Did you have unexplained lapses or convulsions that were believed to represent a seizure or epilepsy? | ☐ | ☐ | ☐ |
| | 30. | **Have any first degree relatives suffered from a seizure, febrile seizure, or epileptic fit?** e.g. Did any of your parents, siblings, or children have unexplained lapses or convulsions that were believed to represent a seizure or epilepsy? | ☐ | ☐ | ☐ |
| | | ☐Mother, ☐Father, ☐Sibling, ☐Child, ☐Other family member:<br><br>**Details:** | | | |
| | 31. | **Did you ever suffer from a migraine headache or its equivalent?** e.g. Did you have unexplained intense stomach aches, severe car sickness, and/or vertigo? | ☐ | ☐ | ☐ |
| | 32. | **Did you have hearing difficulties?** e.g. Did your parents seek out medical attention to evaluate hearing concerns in you? | ☐ | ☐ | ☐ |
| | 33. | **Did you have specific tutoring, or were you in special classes? If yes, please describe what the tutoring was in:** | ☐ | ☐ | ☐ |

| | | | Yes | No | I don't know |
|---|---|---|---|---|---|
| | 34. | **Have you ever been diagnosed with any of the following learning difficulties or disabilities? And if so which one(s):**<br>☐ Dyslexia ☐ Stuttering ☐ Childhood Apraxia of Speech<br>☐ Dyscalculia ☐ Dysgraphia ☐ Dyspraxia<br>☐ Autism ☐ Asperger's ☐ Attention Deficit Hyperactivity Disorder<br>☐ Other: | ☐ | ☐ | ☐ |
| | 35. | **Informally, do you feel that you may have had any of the following learning difficulties or disabilities? And if so which one(s):**<br>☐ Dyslexia ☐ Stuttering ☐ Childhood Apraxia of Speech<br>☐ Dyscalculia ☐ Dysgraphia ☐ Dyspraxia<br>☐ Autism ☐ Asperger's ☐ Attention Deficit Hyperactivity Disorder<br>☐ Other: | ☐ | ☐ | ☐ |
| | 36. | **Have any of your biological parents, siblings, or children been diagnosed with any of the above conditions or have difficulties with any of the above questions from 3-32?**<br><br>**If so, please indicate who and fill in the details below:** | ☐ | ☐ | ☐ |
| | | ☐Mother, ☐Father, ☐Sibling, ☐Child, ☐Other family member:<br><br>**Details:** | | | |
